# Differentiating left ventricular remodelling in aortic stenosis from systemic hypertension and impact of surgical replacement

**DOI:** 10.1101/2023.12.28.23300260

**Authors:** Masliza Mahmod, Kenneth Chan, Joao F. Fernandes, Rina Ariga, Betty Raman, Ernesto Zacur, Ho-fon Royce Law, Marzia Rigolli, Jane Francis, Sairia Dass, Kevin O’Gallagher, Saul Myerson, Theodoros Karamitsos, Stefan Neubauer, Pablo Lamata

## Abstract

(2)

**Background:** Left ventricular (LV) hypertrophy occurs in both aortic stenosis (AS) and systemic hypertension (HTN) in response to wall stress. However, differentiation of hypertrophy due to these two aetiologies is lacking, as well as an understanding of the impact of surgical aortic valve replacement (AVR). The aim was to study the 3-dimensional geometric remodelling pattern in severe AS pre– and post-surgical AVR, and to compare with HTN and healthy controls.

**Methods:** Ninety-one subjects (36 severe AS, 19 HTN and 36 healthy controls) underwent cine cardiac magnetic resonance (CMR). CMR was repeated eight months post-AVR (n=18). Principal component analysis (PCA) was performed on the 3-dimensional meshes reconstructed from 109 CMR myocardial contours of 91 subjects at end-diastole. PCA modes were compared across experimental groups.

**Results:** A unique AS signature was identified by wall thickness linked to a LV left-right axis shift and a decrease in short axis eccentricity. HTN was uniquely linked to increased septal thickness. Combining these three features had good discriminative ability between AS and HTN (AUC=0.792). The LV left-right axis shift was not reversible post-AVR and was predictive of post-operative LV mass regression (R^2^=0.339, p=0.014). AVR was associated with a reduction in global LV size and correction of short axis eccentricity.

**Conclusions:** Unique remodelling signatures can differentiate the aetiology of LV hypertrophy. LV axis shift is characteristic in AS, is not reversible post AVR, predicts mass regression, and is interpreted to be an adaptive mechanism.

Graphical abstract.

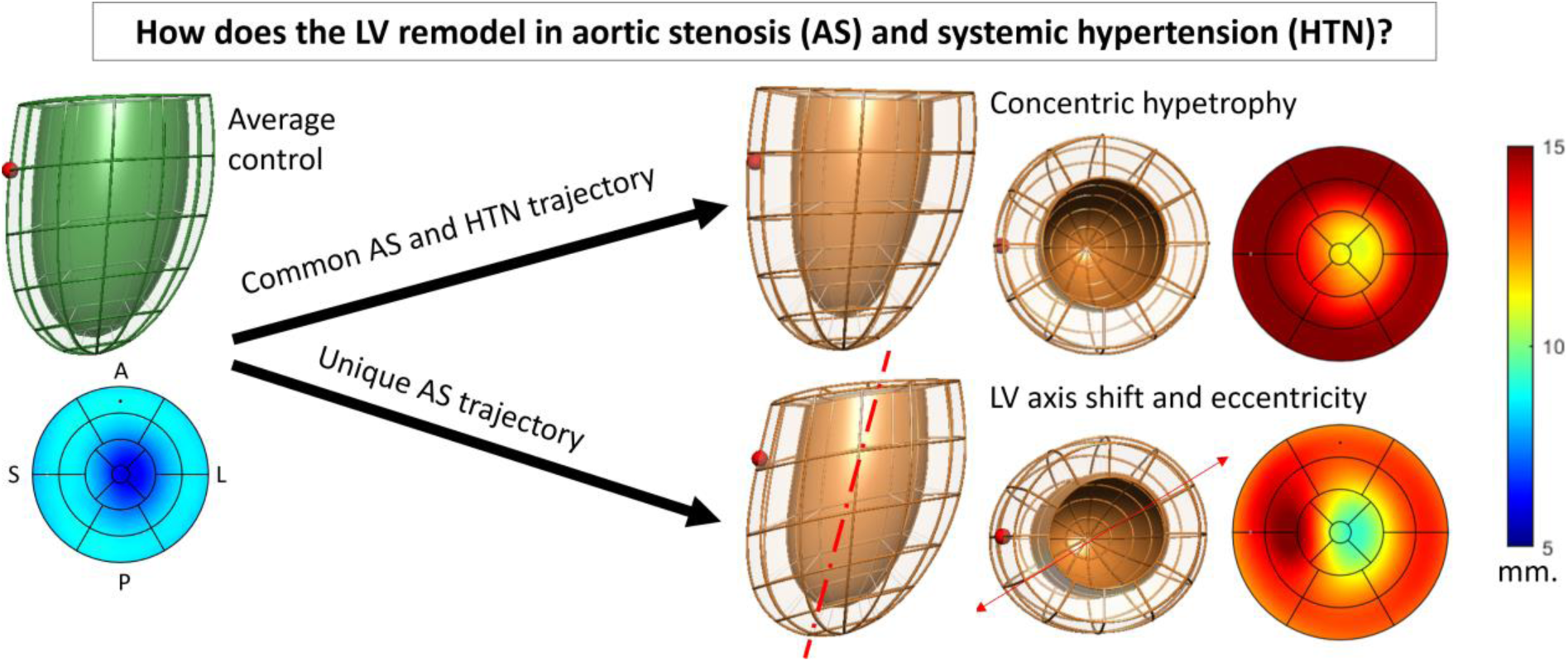

## (3) Introduction

Aortic stenosis (AS) is a common valvular heart disease, occurring in 2% of the population over 65 years of age, rising to 7% in men over 85 years (1). The response of the left ventricle (LV) to AS begins with a compensatory hypertrophic response which serves to normalise LV wall stress. This is characterised by myocyte hypertrophy, thickened LV wall and increased LV mass (2, 3). It ultimately transforms to a maladaptive process, which carries adverse cardiovascular risks, and reversal of this process is accompanied by improvement in outcome (4–6). Surgical aortic valve replacement (AVR) and transcatheter aortic valve replacement are the definitive treatments of symptomatic severe AS, while in asymptomatic severe AS these are recommended if there is LV systolic dysfunction (7).

Systemic hypertension (HTN) presents with a similar pattern of LV remodelling which is mainly concentric LV hypertrophy although eccentric remodelling has been described in about a third of cases (8). While the increased afterload can affect the myocardium in both HTN and AS, the activation of neurohormonal pathways is an additional mechanism that can affect LV remodelling and performance in hypertension (9, 10). HTN is also common in patients with AS and can lead to more severe LV remodelling, premature disease progression and augmented aortic valve calcification (11, 12, 13). When both conditions are present, it is often difficult to differentiate the aetiology of LV hypertrophy in clinical practice. Accurate assessment of LV geometry in AS and HTN could help identify the predominant pathology and therefore guide treatment strategies.

Cardiovascular magnetic resonance (CMR) is an accurate, reproducible and well validated for assessment of cardiac structure and function, and conventional metrics extracted from this imaging modality are mass, volumes and function (14). Recent advances in computational anatomy tools now enable a much more detailed analysis of the LV remodelling through the construction of statistical shape models (SSM, also referred as statistical or computational atlases) of the 3-D geometry of cardiac structures (15). The use of these computational atlases has revealed the impact of a premature birth in the adult heart (16) and has identified a remodelling signature that predicts response to cardiac resynchronization therapy (17).

Using advanced computational anatomy tools, we sought to investigate if: (1) there is a unique remodelling pattern in AS when compared with hypertension; (2) there is an AS remodelling signature that differentiates between symptomatic and asymptomatic patients; and (3) the remodelling pattern associated with AS recovers after AVR.

## Methods

An exploratory study on 109 CMR datasets from 91 participants (18 AS subjects had follow– up data after surgical AVR) was conducted to study the end-diastolic LV morphology with robust and reproducible statistical shape models.

### Study Population

Thirty-six severe AS patients including 26 symptomatic who had NYHA class ≥2 and/or Canadian Cardiac Society Angina, and 10 asymptomatic were prospectively recruited from the Oxford University Hospital National Health Service Trust. Severe AS was diagnosed based on the established criteria (18). All patients had no evidence of significant coronary artery stenosis as shown by invasive coronary angiography. AS patients were included if they had all the criteria of an aortic valve area of ≤1.0 cm^2^, mean pressure drop (PD) of ≥40 mmHg, maximum jet velocity of ≥4.0 m/s and absence of other significant valvular pathology based on clinical echocardiogram. Patients were excluded if they had LVEF <50%, systolic blood pressure BP ≥160 mmHg and diastolic BP ≥90 mmHg, contraindications to MR imaging, glomerular filtration rate <60 ml/min, underlying cardiomyopathy, previous myocardial infarction, coronary revascularization or previous cardiac surgery. Of the 26 symptomatic AS patients who underwent AVR, 18 had a follow-up scan 8 months after AVR. Eight patients did not have a follow-up scan due to perioperative death (2 patients), pacemaker implantation (1 patient), lost to follow-up (1 patient) and did not consent for a repeat CMR (4 patients).

As a comparison, 19 patients with HTN were recruited. All HTN patients had a minimum 2– year history of uncontrolled hypertension with ≥2 antihypertensive agents and had ambulatory BP monitoring ≥140/90 mmHg. In addition, 36 healthy volunteers were recruited as healthy controls – these were asymptomatic, not on medications, had no history of heart disease, diabetes, HTN or high cholesterol and normal physical examination. All subjects gave their informed written consent to participate in the study which was approved by the Institutional Ethics Committee (Ethics number 07/H0607/66).

### Cardiac Magnetic Resonance and Quantification of Ventricular Volumes, Mass and Function

All subjects underwent CMR scanning on a 3 Tesla MR system (TIM Trio; Siemens Healthcare, Erlangen, Germany). Cine imaging was performed using steady-state, free precession breath-hold in long-axis planes and sequential 7-mm short-axis (SAX) slices from the atrioventricular ring to the apex with a 3-mm gap (19). Late gadolinium enhancement (LGE) images were acquired using standard methods as previously described (20). Analysis of cardiac volumes, function and mass was performed according to standard methods. In the AS cohort, aortic valve area was measured from direct planimetry during aortic valve CMR cine. LGE was determined if it was present or not by 2 independent CMR readers (20,21). Two-dimensional feature-tracking (FT) strain analysis was used to derive peak global longitudinal strain from horizontal long-axis cines, while SAX cines were used to derive circumferential and radial strain. All measurements were performed with CMR42 software (version 4.0, Circle Cardiovascular Imaging). Representative images are illustrated in **Figure 1**.

**Figure 1.**
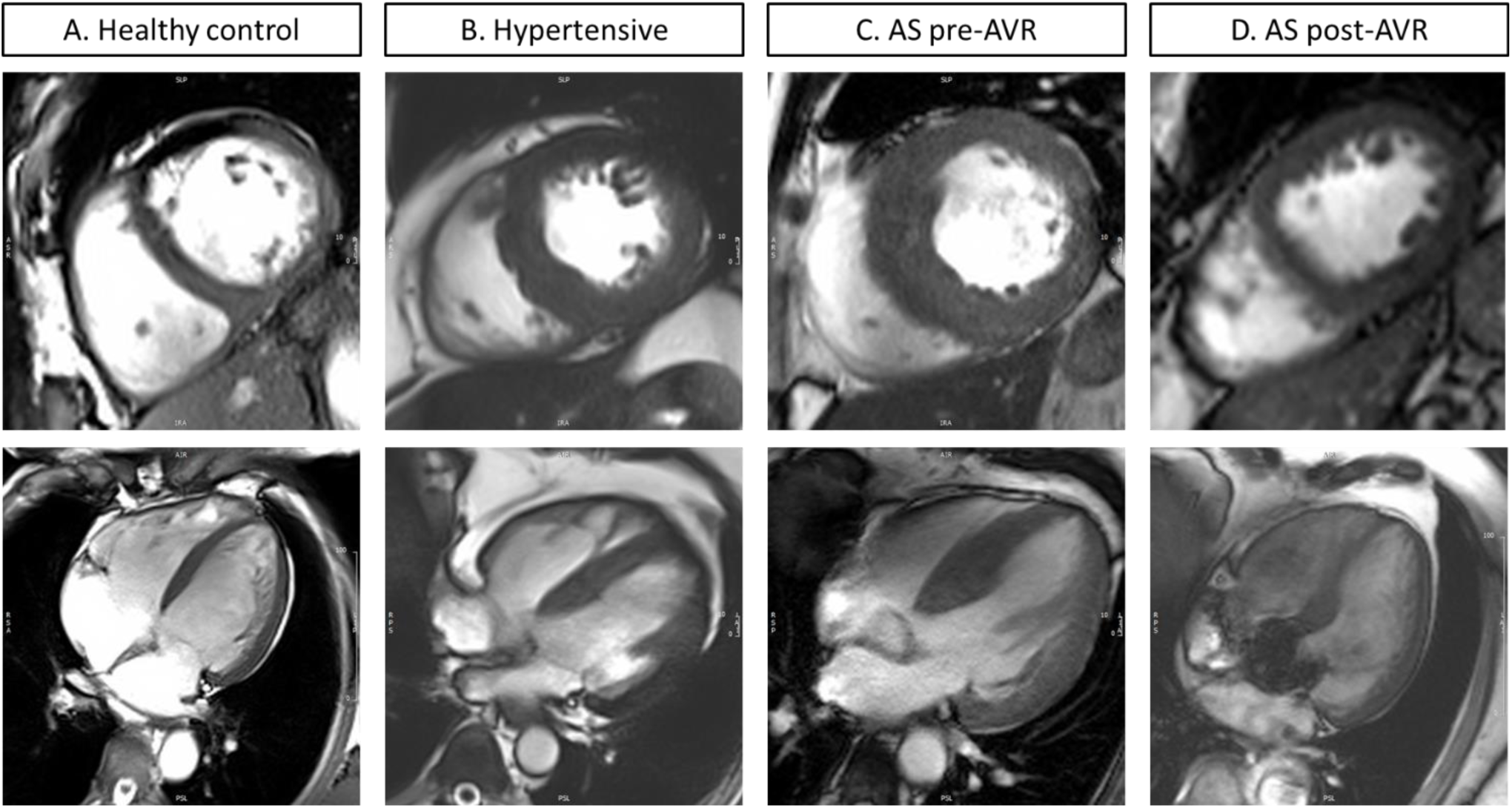
Representative short-axis and 4-chamber view CMR images for. **(A)** healthy control, **(B)** hypertensive patient, **(C)** aortic stenosis pre-AVR, **(D)** post-AVR of the same aortic stenosis patient.

### Statistical Shape Model (SSM) for Assessment of LV Geometry

Creation of an SSM was undertaken using previously published methods (22,23). The end– diastolic frame of the SAX cine stack was manually contoured (LV endocardium and epicardium, RV endocardium) with CVI42 (Circle Cardiovascular Imaging). 3D computational meshes were then fully automatically fitted to contours. The LV mesh of each subject was then described with a mesh defined by a set of 3456 nodal variables (or degrees of freedom).

Once the anatomical information has been captured in these meshes, all 109 shapes were pre– aligned by their centre of mass, by setting the vertical direction as the perpendicular to the SAX slice, and by correction of the relative direction aligning the centre of mass of LV and RV. The average anatomy was found, and a principal component analysis (PCA), a dimensionality reduction technique, was applied to identify the key *PCA modes of anatomical variation* (i.e. the directions of 3D anatomical change that explain the variability of morphologies observed in the 109 shapes). Extreme shapes (±3 standard deviations) of each PCA mode were visually inspected to describe the qualitative shape changes, and the first PCA modes that explained up to 99.5% of shape variability were selected. Each anatomy was now represented as the average anatomy plus the linear combination of the information contained in each PCA mode.

Once the anatomical information is compacted, a supervised learning technique is used to identify the relevant remodelling patterns. The linear combination of the PCA modes that best distinguish between pairs of clinical groups (AS vs. Control; HTN vs. Control; AS vs. HTN; AS vs. AS post-AVR; Asymptomatic AS vs. symptomatic AS) were identified using linear discriminant analysis (LDA). Finally, the LDA shape biomarkers are analysed against clinical parameters using linear regression.

### Statistical analysis

All data are expressed as mean ± standard deviation and checked for normality using Kolmogorov–Smirnov test. Categorical data are presented as numbers and percentages. Comparisons of demographic data, strain and PCA modes between pairs of experimental groups were performed by unpaired t-Test. PCA and LDA were performed with MatLab (The Mathworks, Natick, MA). The generalisability of classification tasks was tested by the area– under curve (AUC) in a leave-one-out cross-validation test as described previously (23).

## Results

### Characteristics of study cohorts

There were no significant differences in age, gender, body mass index (BMI), BP and heart rate between controls and AS. There were no significant differences in gender and BMI between AS and HTN, but HTN subjects were younger (51.4 ± 13.2 vs. 67.6 ± 10.2 years) and had higher BP and heart rate (85.9 ± 16.6 vs 66.8 ± 9.7 bpm) than AS (**Table 1**).

**Table 1:** Baseline characteristics and CMR results of all cohorts. *p value for severe AS vs. normal controls, p value** for severe AS vs. hypertension. SBP, systolic blood pressure; DBP, diastolic blood pressure; LV, left ventricle; LVEDV, LV end-diastolic volume; LVESV, LV end-systolic volume; LVEF, LV ejection fraction; LVMI, LV mass indexed to body surface area; LGE, late gadolinium enhancement; PD, pressure drop.

|  | Severe AS<br>(n = 36) | Normal<br>controls<br>(n = 36) | p-value* | Hypertension<br>(HTN)<br>(n=19) | p-value** |
| --- | --- | --- | --- | --- | --- |
| Age (years) | 67.6 ± 10.2 | 67.7 ± 6.8 | 0.96 | 51.4 ± 13.2 | <0.01 |
| Male, n (%) | 26 (72%) | 19 (53%) | 0.08 | 15 (79%) | 0.75 |
| Body mass index<br>(kg/m<sup>2</sup>) | 27.8 ± 4.6 | 26.2 ± 5.4 | 0.31 | 27.4 ± 6.3 | 0.84 |
| SBP (mmHg) | 134.5 ±<br>16.6 | 126.1 ±<br>13.9 | 0.19 | 149.1 ± 19.9 | 0.007 |
| DBP (mmHg) | 77.0 ± 10.2 | 73.3 ± 9.2 | 0.36 | 86.9 ± 11.7 | 0.002 |
| Heart rate (bpm) | 66.8 ± 9.7 | 61.2 ± 9.3 | 0.11 | 85.9 ± 16.6 | <0.01 |
| Aortic valve PD<br>(mmHg) | 80.1 ± 14.1 | — |  | — |  |
| **Past medical history,**<br>**n (%)** |  |  |  |  |  |
| Hypertension | 11 (31%) | — |  | 19 (100%) | 0.003 |
| Dyslipidaemia | 7 (19%) | — |  | 2 (11%) |  |
| Diabetes | 7 (19%) | — |  | 2 (11%) | 0.17 |
| **Medications, n (%)** |  |  |  |  |  |
| Aspirin | 12 (33%) | — |  | 2 (11%) | 0.02 |
| Metformin | 5 (14%) | — |  | 2 (11%) | 0.22 |
| ACE-I/ARB-II | 9 (25%) | — |  | 10 (53%) | 0.29 |
| Beta-blockers | 7 (19%) | — |  | 7 (37%) | 0.31 |
| Statin | 13 (36%) | — |  | 4 (21%) | 0.04 |
| **CMR results, (± SD)** |  |  |  |  |  |
| LVEDV (ml) | 141 ± 43 | 137 ± 33 | 0.65 | 146 ± 25 | 0.65 |
| LVESV (ml) | 40 ± 21 | 42 ± 12 | 0.62 | 44 ± 15 | 0.53 |
| LVEF (%) | 73 ± 8 | 69 ± 4 | 0.009 | 69 ± 8 | 0.24 |
| LVMI (g/m<sup>2</sup>) | 91 ± 32 | 56 ± 13 | <0.001 | 66 ± 13 | 0.002 |
| Aortic valve area<br>(cm<sup>2</sup>) | 0.85 ± 0.13 | - |  | - |  |
| LGE present, n (%) | 26 (72) | 0 |  | 13 (68) | 0.77 |

Of the 26 symptomatic AS patients, 18 (69%) had dyspnoea, 8 (31%) had angina and 1 (<1%) had syncope. CMR results revealed that AS patients had increased LVMI when compared to HTN and control (**Table 1**). Global LV longitudinal strain was lowest in HTN when compared to AS and controls, while global circumferential strain was lower in AS and HTN when compared to controls (**Table 2**).

**Table 2.**
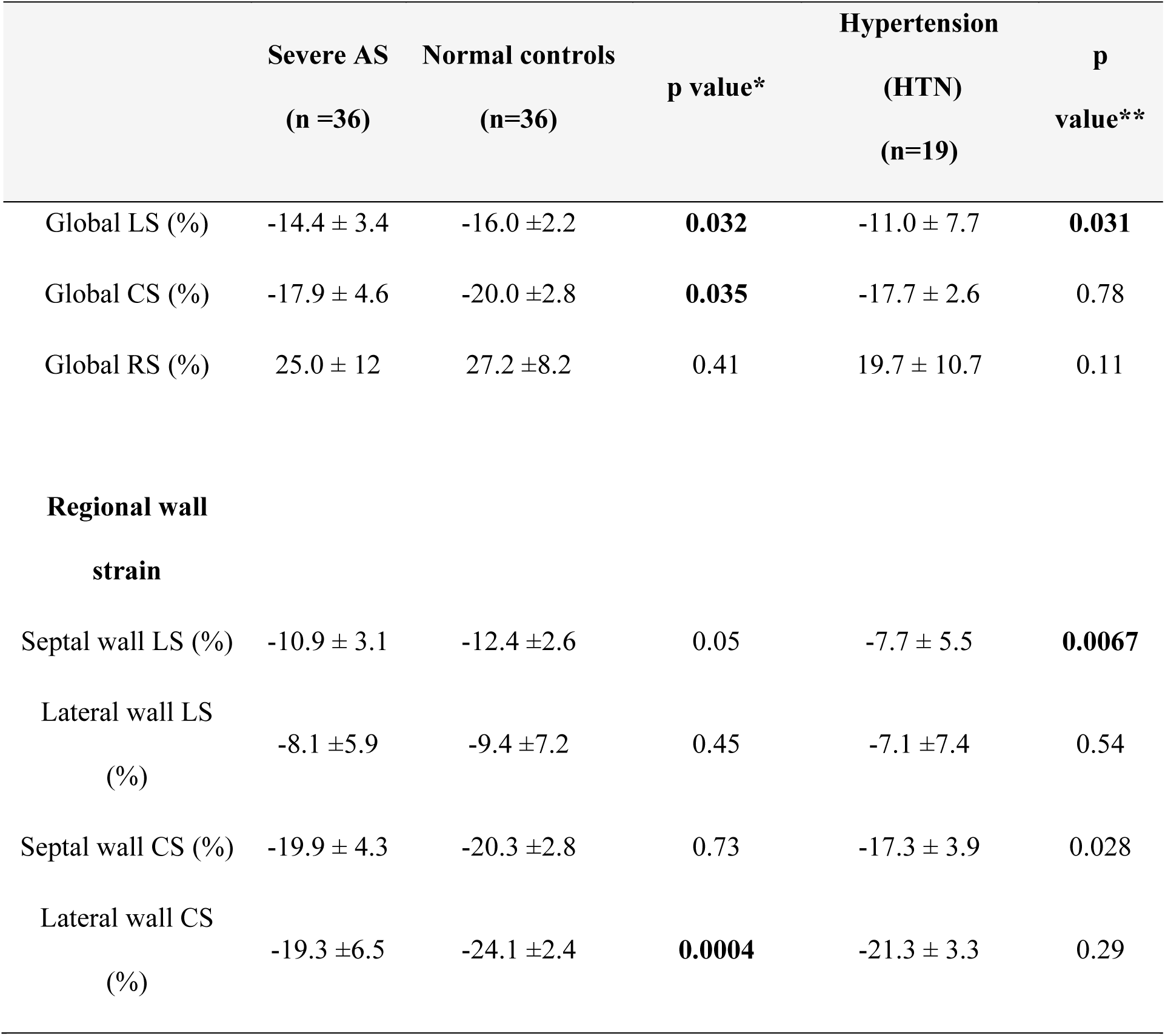
Feature tracking of left ventricular strain. Regional wall defined by American Heart Association (AHA) 17-segment model. (CS, circumferential strain; LS, longitudinal strain; RS, radial strain) *p value for severe AS vs. normal controls, p value** severe AS vs. hypertension.

### Mesh fitting accuracy and pattern of geometric shapes

The LV mesh derived from the contours achieved an excellent sub-voxel accuracy with average fitting error of 1.24 mm (**Figure 2.A**). The first 18 PCA modes of variance accounted for 99.56% cumulative variance captured in the 109 meshes (**Figure 2.B**) over the average LV 3D shape illustrated in **Figure 2.C**. The modes that captured differences are illustrated in **Figure 2.D**.

**Figure 2.**
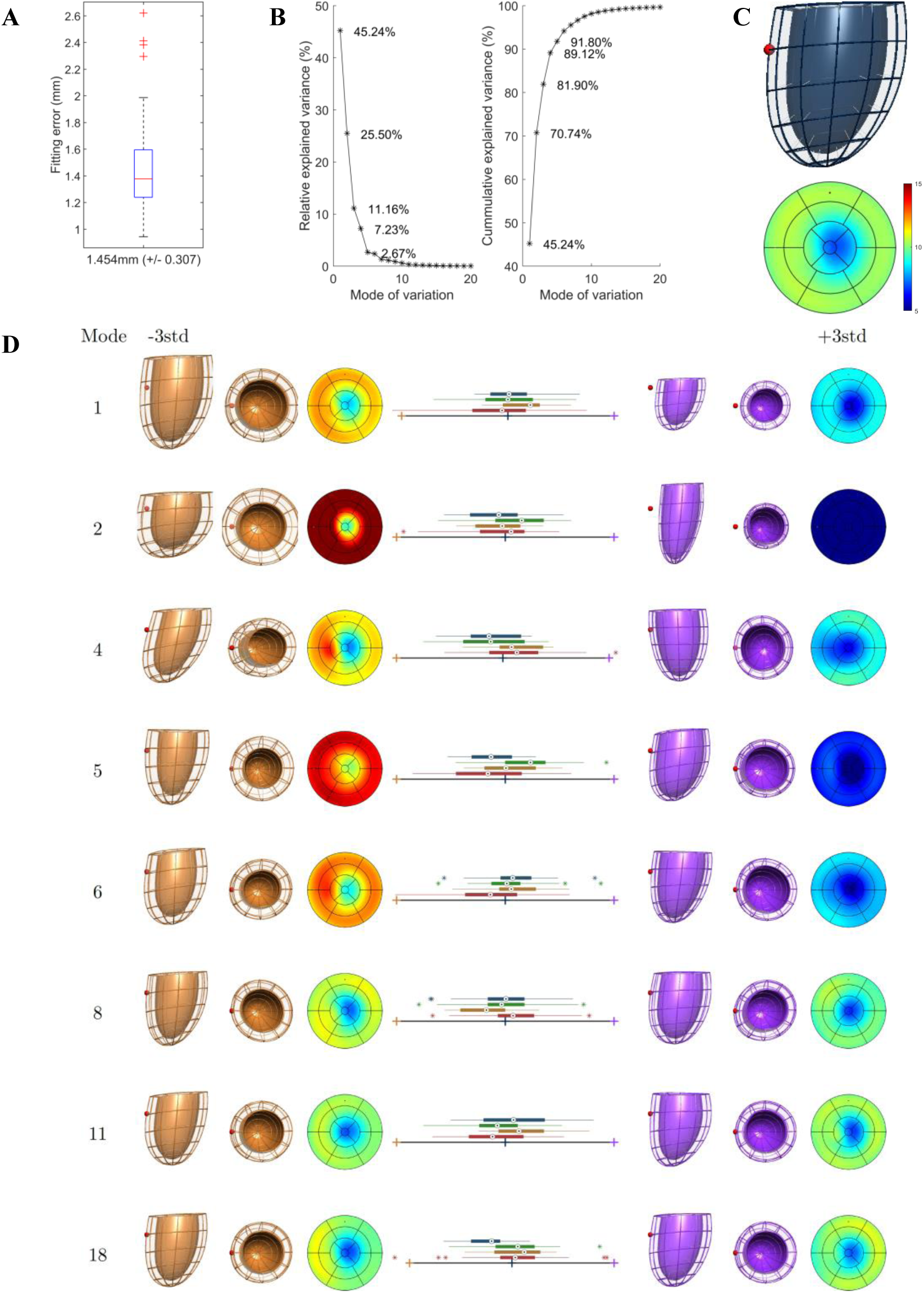
Statistical shape model of the LV anatomy of our cohort. **(A)** Geometrical fitting error of the 109 meshes; **(B)** Individual and cumulative variance in LV shape explained by the PCA modes; **(C)** Anterior view of the average 3D shape (red sphere located in septal wall) and its corresponding thickness bullseye plot (in mm); **(D)** Extreme shapes (+/– 3std) encoded in each PCA mode, with box-plots of the four experimental groups (AS in red, n=36; AS after AVR in orange, n=18; controls in green, n=36; HTN in blue, n=19).

### Comparison of geometric changes in AS, HTN, healthy controls, and post-AVR

Four group comparisons were made. Common and unique discriminative features found are summarised in **Table 3** and illustrated in **Figure 3**.

**Figure 3.**
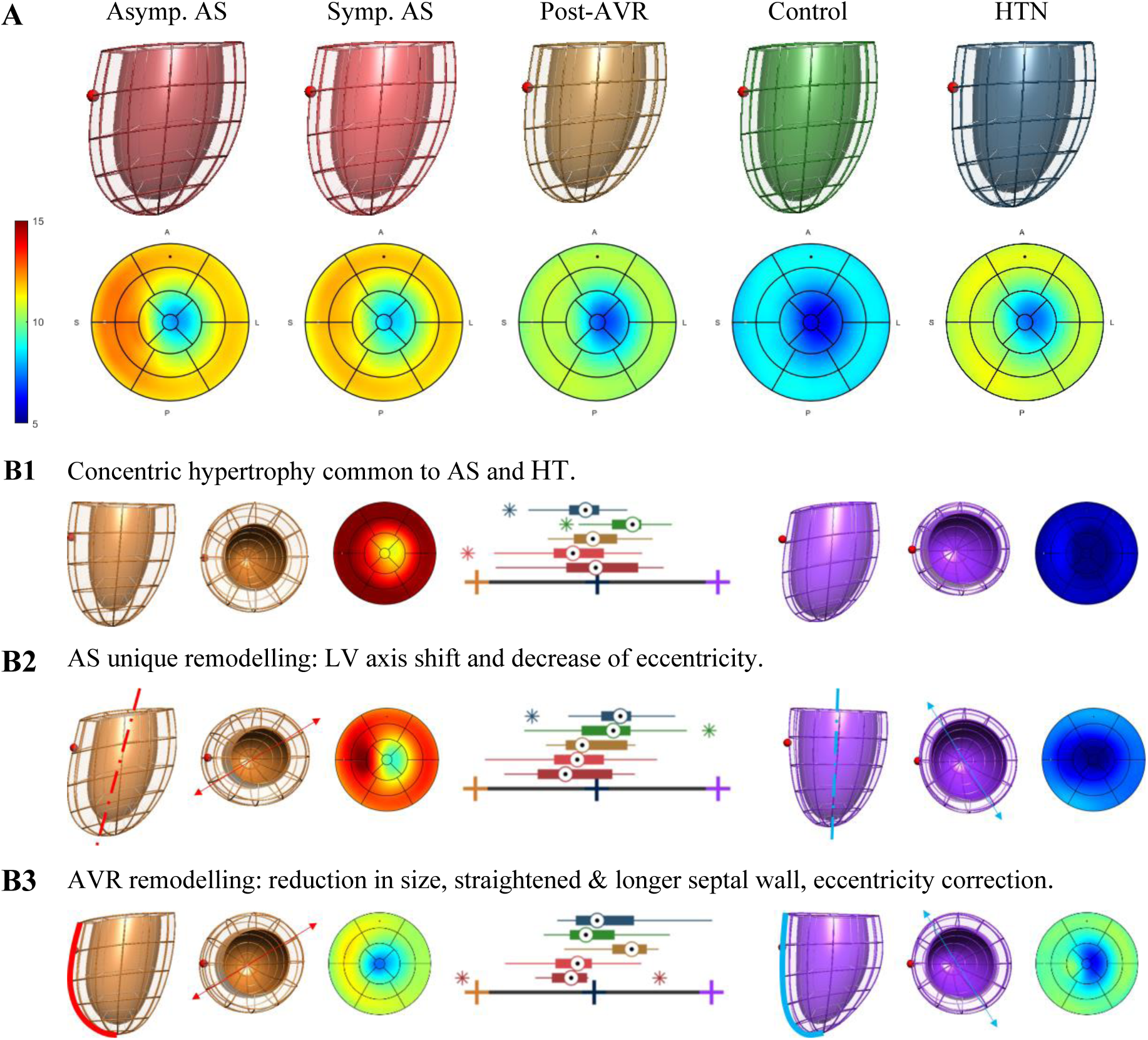
Average shapes and discriminative axes of remodelling. **(A)** Average anatomies and corresponding thickness bullseye plot (in mm) of asymptomatic severe AS before AVR (dark red, n=18), symptomatic (light red, n=18), post AVR (gold, n=18), controls (green, n=36) and HTN (blue, n=19). The red sphere indicates the location of the right ventricle. **(B1)** The axis that captured the common traits of AS and HTN vs. controls (i.e. LDA from modes 2 and 5). **(B2)** The axis that captured the unique traits of AS vs. controls and HTN (i.e. LDA from modes 4 and 6). **(B1)** The axis that captured the impact of AVR (i.e. LDA from modes 1, 5, 6, 8 and 11).

**Table 3.**
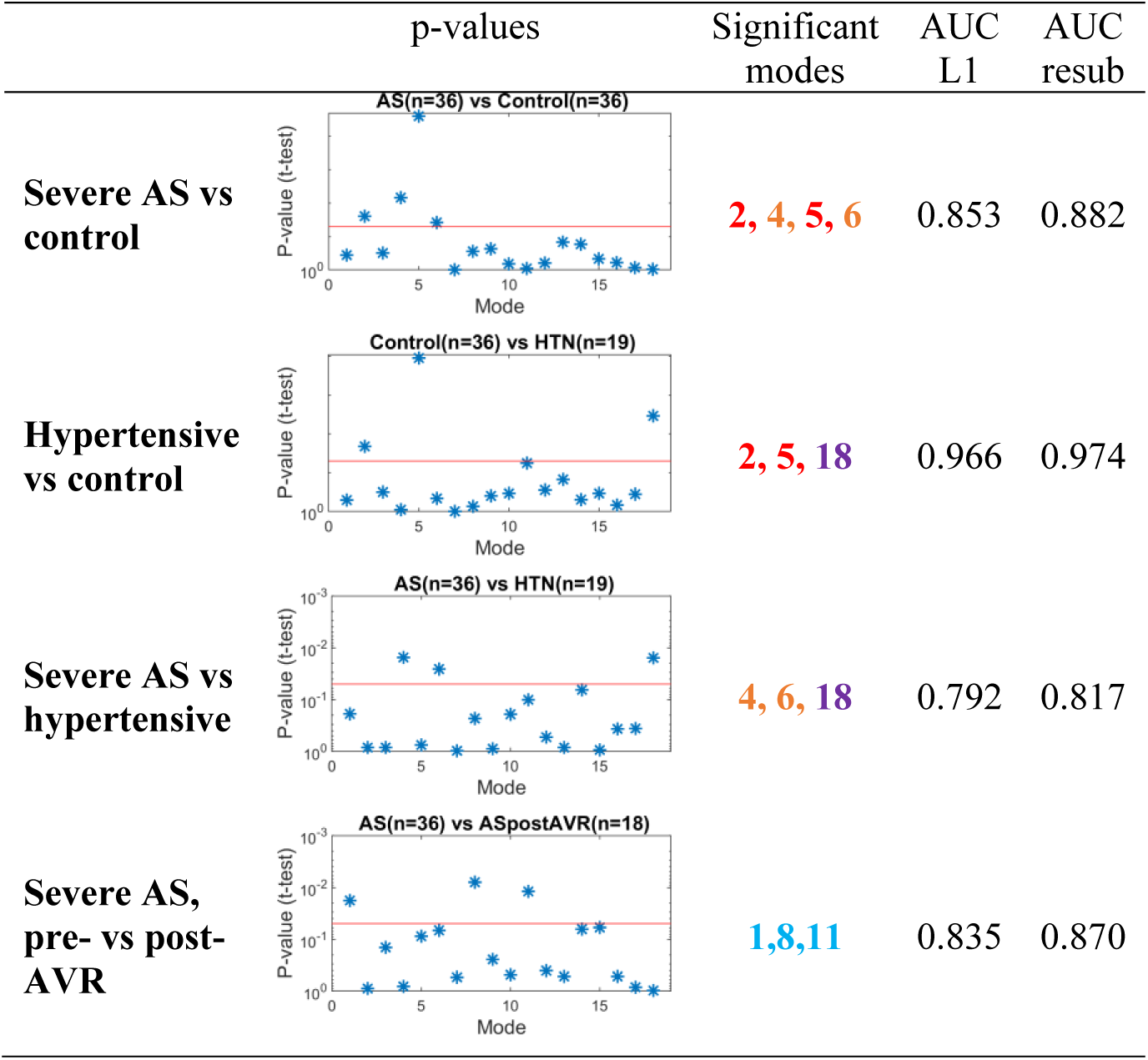
Identification of PCA modes that are significant (p<0.05) in differentiating between experimental groups. The signature of an increased afterload are modes **2** and **5** (common in AS and HTN when compared to controls). The unique signature of AS is mode **4** and **6** (both differentiate AS to HYT and controls). The unique signature of HTN is mode **18**. The signature of correction of AS by AVR is mode **1**, **8** and **11**. See Figure 2 for an illustration and interpretation of these modes.

|  | p-values | Significant modes | AUC L1 | AUC resub |
| --- | --- | --- | --- | --- |
| **Severe AS vs control** |  | **2, 4, 5, 6** | 0.853 | 0.882 |
| **Hypertensive vs control** |  | **2, 5, 18** | 0.966 | 0.974 |
| **Severe AS vs hypertensive** |  | **4, 6, 18** | 0.792 | 0.817 |
| **Severe AS, pre- vs post-AVR** |  | **1, 8, 11** | 0.835 | 0.870 |

The common impact of HTN and AS in LV anatomy was the development of LV hypertrophy, as shown by the combination of PCA mode 2 (reduced LV length and increased LV wall thickness) and PCA mode 5 (concentric hypertrophy). Mode 5 was the feature that best differentiated controls from both AS and HTN.

In addition to these common characteristics, two unique PCA modes that differentiated AS from both controls and HTN were found: AS displayed an LV axis shift (apex shifts to the right, mode 4) as a predominant feature, combined with an outward remodelling of the outflow track (OT) of mode 6. Both modes also displayed increased wall thickness and a decrease in short axis eccentricity, i.e. the ratio between the left-right to the anterior-posterior diameter (24), linked to AS. Combining the significant modes 2, 4, 5 and 6, an excellent discriminatory performance was reached between AS and controls (leave-one-out-AUC = 0.853).

HTN was also found to have a unique characteristic that differentiated it from the AS and control groups: the presence of a mild septal hypertrophy as captured by PCA mode 18. HTN shape by modes 2, 5 and 18 reached an outstanding discriminative ability between HTN and controls (leave-one-out-AUC = 0.966).

The discrimination between AS and HTN was the most challenging task: combination of the significant PCA modes 4, 6 and 18 led to a good performance (leave-one-out-AUC = 0.792).

Finally, AVR resulted in a reduction in LV size (mode 1), correction of short axis eccentricity (mode 8) and a longer and straightened septal wall (mode 11), reaching an excellent discriminative ability between AS pre– and post– AVR (leave-one-out-AUC = 0.835).

Although not reaching statistical significance, AVR showed the tendency of reduction in LV wall thickness, both independent (mode 5) and associated with short axis eccentricity (mode 6). There was no correction for mode 2 (thicker walls and shorter LV) and mode 4 (LV axis shift) detected.

### Comparison of geometric changes between symptomatic & asymptomatic AS

No differences were found in any of the 3 axes of remodelling (the concentric remodelling common to AS and HT, the unique AS remodelling, and the AVR remodelling, see panels B1-B3 in Figure 3) between the symptomatic and asymptomatic AS subgroups. In the study of the individual PCA modes, the asymptomatic have less concentric hypertrophy accordingly to mode 5 (p=0.026), and a tendency of more concentric hypertrophy and increased sphericity accordingly to mode 2 (p=0.18), with a net effect of no change in thickness or mass but an increase in sphericity.

### Correlations between clinical parameters before and after AVR

At baseline prior to AVR we investigated the link between LV morphology and LV function. LV thickness and length (modes 1 and 2) had good correlations with global systolic strain (p=0.0007, p=0.0003 and p=0.003 for radial, circumferential, and longitudinal strain respectively) (**Table 4**). A linear combination of modes capturing LV axis shift and outflow tract remodelling (modes 3,4,6,7,8) was best associated with baseline aortic pressure drop (PD) (**Table 4**).

**Table 4.** Correlations between shape biomarkers (i.e. LDA combination of PCA modes) and clinical parameters pre-and post AVR. AVR, aortic valve replacement; LV, left ventricle; PD, pressure drop.

| Clinical parameter | PCA<br>mode | Qualitative description | R<sup>2</sup> | p-value |
| --- | --- | --- | --- | --- |
| **Pre-AVR** |  |  |  |  |
| LV strain |  |  |  |  |
| Radial strain | 1,2 | LV size and length (sphericity) | 0.22 | 0.004 |
| Circumferential strain | 1,2 | LV size and length (sphericity) | 0.27 | 0.001 |
| Longitudinal strain | 1,2 | LV size and length (sphericity) | 0.19 | 0.009 |
| Aortic PD pre-AVR | 3,4,6,7,8 | LV axis and outflow tract<br>remodeling | 0.13 | 0.008 |
| **Post-AVR** |  |  |  |  |
| Aortic PD post-AVR | 1,5,6,7,8 | Ventricular size, sphericity and<br>outflow tract remodeling | 0.47 | 0.002 |
| Reduction in aortic PD | 1,5,6,7,8 | Ventricular size, sphericity and<br>outflow tract remodeling | 0.53 | <0.001 |
| %mass regression | 4,6 | LV axis | 0.34 | 0.014 |

We also investigated the ability of anatomical biomarkers to predict the outcomes of AVR. Baseline ventricular thickness, cavity sphericity and outflow tract remodelling (modes 1,5,6,7 and 8) were best correlated with aortic PD after AVR (p=0.006) as well as the absolute PD reduction (p=0.0017). However, this remodelling pattern was not predictive of the percentage of mass regression at 8 months follow-up (R=0.16, p=0.11). Instead, the unique AS signature of LV axis shift (mode 4) correlated with mass regression (R^2^=0.339, p=0.014).

## Discussion

We found a unique LV hypertrophy remodelling pattern in severe AS that is distinctive from systemic HTN: severe AS displayed an LV axis shift that is observed in both asymptomatic and symptomatic subgroups. For symptomatic AS patients who underwent AVR, the LV axis shift did not change significantly in the post-operative follow-up CMR but was associated with a greater degree of LV mass regression, suggesting that LV axis shift could be an adaptive remodelling pattern in response to the stenotic aortic valve.

### A unique LV remodelling differentiating severe AS and HTN

In patients with both AS and HTN, distinguishing AS from hypertensive LV hypertrophy remains challenging. HTN (i.e. reduction in systemic arterial compliance and/or increase in vascular resistance) and AS (i.e. proximal increase of resistance to the LV outflow) can cause different types of pressure overload, and we therefore hypothesize that they should also lead to different remodelling patterns beyond the primarily concentric LV hypertrophy (25).

It has been shown that in AS patients with concomitant hypertension, symptoms develop early with a relatively larger aortic valve area and lower stroke work loss when compared to patients with isolated AS, which suggests tight control of BP in these patients should be achieved (25). Understanding the different contributions of AS and hypertension on ventricular remodelling could facilitate personalised treatment strategy for patients, whether for intervention to relieve valve stenosis or intensify pharmacological treatments for hypertension.

The LV axis shift (i.e. mode 4 in **Figure 2.D**) is the most distinctive remodelling pattern found to be specific to AS when compared to the pattern seen in HTN, and is a novel feature of our cohort. Similar LV axis shifts have been previously reported to be associated with preterm birth in young adults, i.e. PCA mode 4 in Lewandowski et al (16), and obesity in children, i.e. PCA mode 3 in Marciniak et al (26). The early stage of cardiac adaptation in preterm birth and infant obesity suggests that the axis shift is a trait of adaptive remodelling.

In comparison to these previous studies, PCA mode 4 in severe AS also captures a localised increased thickness of the septal wall, indicating the presence of an elevated work/stress in this part of the myocardium linked to the axis shift. Our HTN subjects did also have the feature of thickened septal walls, but milder in magnitude and independent of the LV axis shift (compare mode 18 to mode 4 in **Figure2.D**). Laplace’s Law explains that the flatter the surface, the larger the stress, and it may be the mechanism of the development of these thickened septal walls (since the septum presents a flatter curvature) as in the onset of basal septal hypertrophy in HTN (27).

The balance of the workload between the septal and lateral walls is a likely associated mechanistic link to explain localised thickening patterns: whereas our HTN patients displayed an impaired septal longitudinal strain in agreement with previous studies (27), our AS patients presented with an impaired lateral wall circumferential strain (see **Table 2**). The ability to assess regional indexed myocardial work has revealed progressive imbalance of the distribution of workload in HTN, with a smaller load in the basal regions with respect to the apical regions (27), and is thus an opportunity for further research into these mechanistic links.

This axis shift must be interpreted with respect to the pre-alignment convention taken when building the Statistical Shape Model, i.e. the vertical direction was defined in this and previous studies (16, 26) as the perpendicular to the short axis MRI plane. The actual 3D re– orientation is illustrated in **Figure 4**. This is interpreted as the result of the interplay between the elongation of the aorta causing a downward shift of the aortic root, hence increasing the angle between LVOT and the aorta in AS (28), and an increase in the local afterload of the stenotic valve (i.e. valve acting as a rigid structure in the heart), confined within the fixed pericardial space (which allows the basal plane to be shifted).

**Figure 4.**
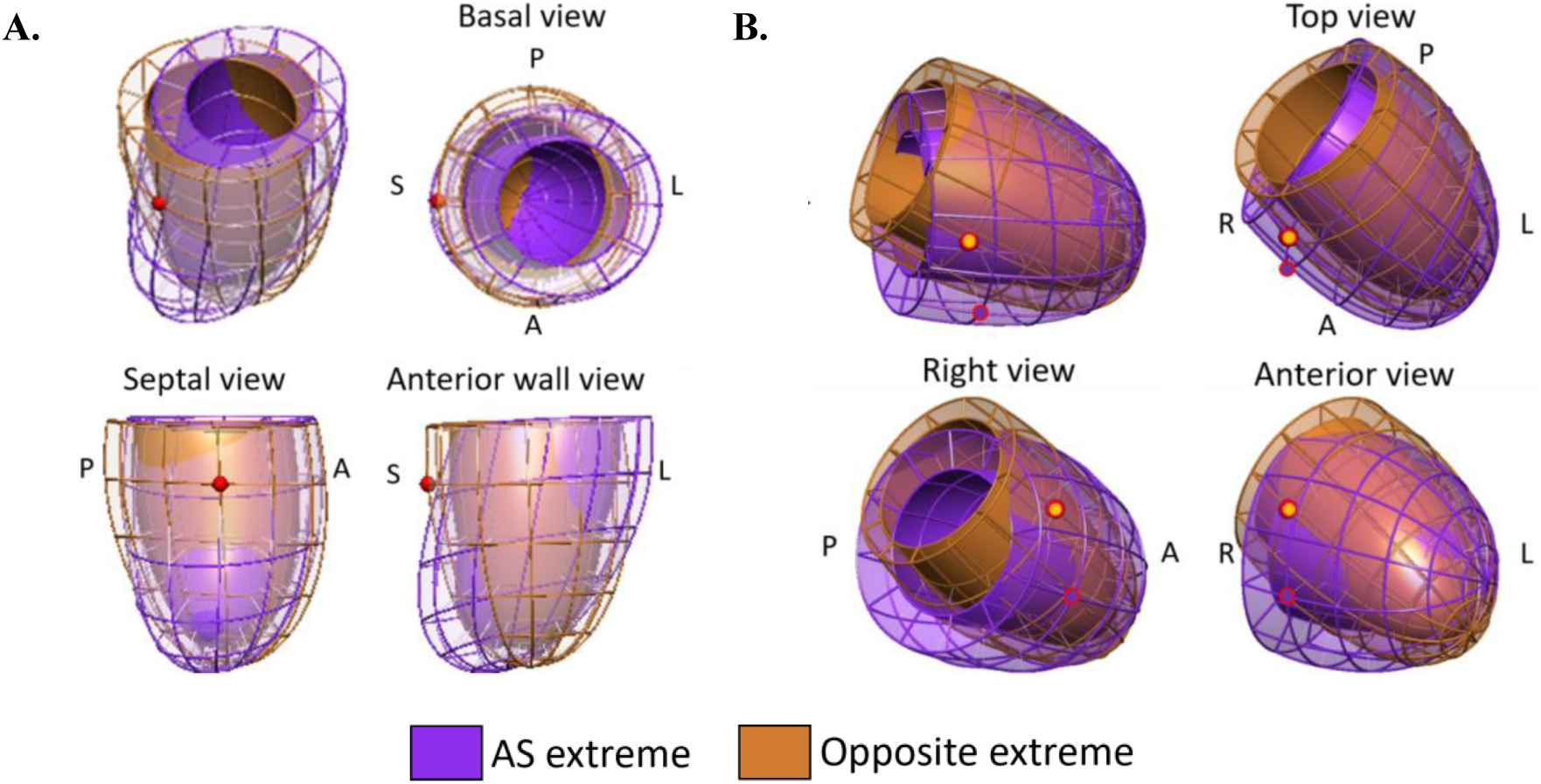
The impact of AS in LV morphology in two systems of coordinates, illustrated as the overlap between the extreme shapes that maximises the differences between controls and AS. **Panel A**: Results generated in heart’s local coordinate system (i.e. LV shapes aligned by the basal plane, and by the direction joining the centre of mass of LV and RV), equivalent to the combination of features illustrated in Figure 2. Note that the red sphere shows the consistent location of the septal wall in both extremes. **Panel B**: Results found in a SSM built in patient’s coordinate system, where the LV shapes are pre-aligned by their centre of mass only. The gold and velvet spheres indicate the different location of the septal wall in each extreme.

We have also identified the known features of ventricular remodelling pattern in AS that share similarities with HTN such as bulk concentric remodelling (i.e. increased ventricular wall thickness captured by modes 2 and 5 in **Figure 2.D**). As expected, the degree of LV hypertrophy was higher in the severe AS group given only mildly elevated BP in the hypertensive group. AS and HTN commonly coexist as shown by a third (31%) of our AS cohort had concomitant HTN, in line with a previous study (25). Given the well-controlled BP in the hypertensive AS suggests that the LV axis shift was predominantly due to AS.

### The interplay between LV anatomy and function

In a search for further evidence of the adaptive vs. maladaptive nature of the changes in LV morphology, we sought to study the interplay between these changes and LV function. The most relevant finding was that only the bulk changes in size (PCA modes 1 and 2 that explain most of the changes in mass, length, volume or sphericity) correlated with the changes in all 3 global strains. On the contrary, the stenotic burden (i.e. the pressure drop) correlated with a certain combination of other morphological features such as the LV axis shift (see **Table 4**). LV morphology is accepted to be a contributing factor to the obstruction in hypertrophic cardiomyopathy (29), but in AS this causal link is not very plausible, and the correlation between LV morphology and stenotic burden is interpreted as an additional evidence that the LV adapts to the presence of AS in unique ways beyond hypertrophy.

### Prediction of AVR outcomes and understanding the impact of AVR

In patients with severe symptomatic AS who had AVR, the LV shift remodelling did not change significantly postoperatively after 8 months and was a feature predictive of LV mass regression. We interpret this finding as another supporting evidence to define the LV shift as a trait of an adaptive remodelling, i.e. as a compensatory mechanism that heralds the ability to further recover (i.e. regress mass) after AVR. It is well established that LV mass regression after aortic valve intervention predicts favourable clinical outcomes such as reduction in heart failure and mortality (30).

AVR was also associated with a reduction of LV size (mode 1, **Figure 1** **panel D**) when compared to pre-AVR subjects, a different feature compared to the concentric hypertrophy characteristic of HTN and severe AS (modes 2 and 5, **Figure 3** **panel B1**), and a correction of short axis eccentricity and a straightened and longer septal wall (modes 8 and 11, **Figure 3** **panel B3**). This finding suggests that there are two different remodelling trajectories, a concentric remodelling when the heart gradually adapts to the growing burden caused by a stenotic valve, and the aforementioned characteristics when the heart has a sudden relief of that burden after replacement of the valve. The lack of complete reversal of LV concentric hypertrophy at 8 months may merely indicate incomplete LV recovery as it has been shown that LV mass regression can occur up to 2 years (31). Therefore, future studies evaluating the long-term effect of AVR on LV geometry may be informative.

### Asymptomatic and symptomatic severe AS have similar remodelling

The current study found no differences in the main 3 axis of remodelling. There was a tendency of symptomatic subgroup to display an increased concentric hypertrophy whilst asymptomatic subgroup to display a larger axis shift (see Figure 3, panels B1-B3), although these were not statistically significant. The interpretation of the LV axis shift as an adaptive mechanism suggests the hypothesis that the lack of this anatomical trait in severe AS could be detrimental, as it may indicate the inability of the heart to compensate for the extra burden caused by the stenotic valve. The preliminary supportive evidence provided here is the ability of this anatomical trait to predict mass regression. Future studies are warranted to contrast this hypothesis, and to study the remodelling trajectories in AS (i.e. mild and moderate severities), and the interplay between 3D macro-remodelling and micro-remodelling (e.g. fibrosis revealed by late gadolinium enhancement) and function, while dissecting gender (e.g. male heart develops an accentuated hypertrophic phenotype (33)) and ethnic differences.

### Limitations

There are several limitations of our study. Firstly, the variations in AS shape remodelling are dependent on the total number of hearts analysed, thus a larger pool of sample will considerably increase the generalisability of the results and enable the analysis of smaller modes of anatomical variation that encode for subtle hypertrophy patterns.

Secondly, since our aim was to study hypertensive patients without other comorbidities, our hypertensive patient cohort were understandably younger than AS patients. To mitigate this limitation, two further sub-analysis were done: first, a subgroup comparison of age– matched patients (AS group n=30, mean age 67.9±8.4; hypertensive group n=9, mean age 62.4±2.7; p=0.06) was performed and showed minimal impact on the discriminative power between the 2 groups (AUC 0.719 vs 0.720 with whole cohort). Second, a correlation analysis between age and the PCA modes only revealed mode 8 as significant (p<0.01), a mode that is not involved in the differences between AS and HTM groups.

Lastly, the surgical outcome was assessed with follow-up CMR at 8 months. Further prospective study could specifically investigate the long-term prognostic significance of different pattern of ventricular remodelling, and differences between AVR and the transcatheter approach.

## Conclusions

Severe AS is characterised by unique LV remodelling patterns when compared to HTN. The LV axis shift is a remodelling trait interpreted as adaptive, that is associated with mass regression, and that might be a potential marker for personalised risk stratification in the management of AS.

**Abbreviations:** AUC, area-under-curve; AS, aortic stenosis; AVR, aortic valve replacement; BP, blood pressure; CMR, cardiac magnetic resonance; HTN, systemic hypertension; LDA, linear discrimination analysis; LGE, late gadolinium enhancement; LV, left ventricle; NYHA, New York Heart Association; PCA, principle component analysis; PD, pressure drop; RV, right ventricle; SAX, short-axis; SSM, Statistical Shape Model.

## Data Availability

The data underlying this article are available in FigShare, at 10.6084/m9.figshare.23815770. Reviewers can access the contents of this data before it is released at https://figshare.com/s/a867a8edb01ba2596a3e

## (4) Acknowledgments

Authors thank Michelle D’Souza for her contribution in the segmentation task of this work.

## (5) Sources of funding

This work was supported by BHF Translational Award (TG/17/3/33406); EU’s Horizon 2020 R&I programme under the Marie Skłodowska-Curie (764738); National Institute of Health Research (NIHR) Academic Clinical Fellowship (KC); Wellcome/EPSRC Centre for Medical Engineering (WT203148/Z/16/Z); Wellcome Trust Senior Research Fellowship (209450/Z/17/Z) (PL). SGM acknowledges funding support from the Oxford NIHR (National Institute for Health Research) Biomedical Research Centre.

## (6) Disclosures

n.a.

## Notes

### Competing Interest Statement

The authors have declared no competing interest.

### Author Declarations

All subjects gave their informed written consent to participate in the study which was approved by Buchkinghamshire, England Research Ethics Committee (Ethics number 07/H0607/66).

